# Socioeconomic and sociodemographic inequalities in the continuum of maternal healthcare utilization in Kenya: Evidence from the 2022 Kenya Demographic and Health Survey

**DOI:** 10.64898/2026.06.24.26356459

**Authors:** Nita Claris Akech, Ken Marvel Lubongah, Philip Okothe Ojuola

## Abstract

**Background:** The continuum of maternal healthcare utilization is crucial for enhancing maternal and child health outcomes. Despite high individual service utilization, completion of the continuum of care among many women in Kenya remains suboptimal, with significant attrition across the stages of care. This study examined socioeconomic and sociodemographic inequalities in the continuum of maternal healthcare utilization in Kenya.

**Methods:** This study used the 2022 Kenya Demographic and Health Survey, specifically the women’s fileA total of 13,612 women aged 15-49 years who had a live birth within 5 years before the survey were included in the analysis. Continuum of care was categorized as no, partial, or complete. A multivariable multinomial logistic regression model was used to estimate inequalities across the continuum of care, with no continuum as the reference category. Results were expressed as adjusted Relative Risk Ratios (aRRR) with 95% confidence intervals.

**Results:** Overall, approximately 36.6% (95% CI: 35.1-38.0) achieved a complete continuum, while a majority, 60.8% (95% CI: 59.3 – 62.2), achieved a partial continuum. In terms of individual service utilization, 67% of women had 4 or more antenatal care contacts, 62% had a skilled delivery, and 54% had a postnatal visit within 48 hours of delivery. We observed inequalities across parity, age, wealth quintiles, education, pregnancy intention, place of residence, and media exposure that influenced both the partial and complete continuum of maternal healthcare utilization.

**Conclusion:** Approximately one-third of women in Kenya received a complete continuum of care during the prenatal and postnatal periods. The government and stakeholders should prioritize targeted, equity-focused interventions for women with no education and those from low-income households to increase service utilization. Non-governmental organizations and the Ministry of Health should also intensify media sensitization and expand access to family planning services.

**Contributions to Literature:**

- There is limited literature in Kenya assessing the full spectrum of maternal healthcare utilization or the inequalities within the Continuum of care. Most studies examine individual service utilization, which can obscure critical gaps in care.
- This study aims to assess the socioeconomic and sociodemographic inequalities in the continuum of maternal healthcare utilization by utilizing the nationally representative KDHS.
- The findings of this study will help develop interventions and policies aimed at bridging the gaps in maternal service utilization, thus improving maternal and child health outcomes in Kenya.

## Introduction

Maternal mortality is a key indicator of health system performance and equity and a major global health concern. Despite a 40% decline in maternal mortality between 2000 and 2023[1], an estimated 260,000 maternal deaths still occur annually. A majority of these deaths (70%) occur in low- to middle-income countries (LMICs), particularly in Sub-Saharan Africa (SSA) [1]. These deaths are preventable with access to essential maternal healthcare services, including antenatal care (ANC), skilled birth attendance (SBA), and postnatal care (PNC). In recent years, attention has shifted from individual service utilization to the continuum of care, which emphasizes sustained engagement with maternal healthcare services during the prenatal, childbirth, and postnatal periods[2–4].

Although global improvements in service utilization have been observed, substantial inequalities persist across the continuum of care. While many women attend at least one ANC visit, very few complete the recommended number of ANC visits[5], deliver with skilled attendance, and receive timely postnatal care[6]. These drop-offs are critical because failure to achieve a complete continuum is associated with adverse maternal and neonatal outcomes[7–9]. The continuum of care framework provides a comprehensive approach to assessing maternal healthcare system performance and identifying service delivery gaps[4].

SSA has seen uneven progress in maternal healthcare utilization. The region continues to account for the largest proportion of maternal deaths globally[1, 10]. Completion of the maternal healthcare continuum remains low, with persistent inequalities driven by socioeconomic status, geographic access, and education [4, 11, 12]. Women from poorer households, those residing in rural areas[12] and those with low educational attainment[13] are less likely to complete the continuum of care. This highlights persistent health system and structural barriers[14] underscoring the need for equity-focused interventions to improve maternal health service delivery.

Over the years, maternal health outcomes in Kenya have improved; however, significant gaps remain in ensuring continuity of care. According to national data, there is a high rate of initial ANC contact, with substantial attrition across subsequent stages of care, particularly in postnatal service utilization [15, 16]. Recent studies in Kenya that have utilized DHS data focus on antenatal care, skilled delivery, or postnatal service utilization in isolation rather than on the full continuum of care or the inequalities within it, potentially obscuring critical gaps in continuity. We examined socioeconomic and sociodemographic inequalities across the continuum of maternal healthcare utilization in Kenya using nationally representative data from the Demographic and Health Survey (DHS). The findings of this study will provide a better understanding of maternal healthcare utilization patterns and identify populations at risk of discontinuity in Kenya. Additionally, they will help formulate interventions and policies targeted towards improving maternal health outcomes in Kenya.

## Methods

### Data Source and Population

The study used secondary data from the 2022 KDHS. The KDHS is conducted periodically to collect data on key population health indicators, including maternal and child health indicators [15]. The women’s file (IR) was used for this study. We accessed the data on Jun 8, 2026, from the DHS program after obtaining approval. We did not access any personally identifiable information, as it had already been redacted by the time the data was being accessed. The analysis was restricted to women aged 15-49 years who had at least one live birth in the five years preceding the survey. In addition, only women whose most recent child was less than 60 months old at the time of the survey were included to assess the completeness of maternal healthcare utilization. A total weighted sample of 13,612 women was used for the analysis.

### Outcome Variable: Continuum of Maternal Healthcare Utilization

The primary outcome was the continuum of maternal healthcare utilization, defined as receipt of three essential services: four or more ANC contacts, a skilled delivery, and postnatal care (PNC) within 48 hours of delivery. We defined adequate ANC as four or more ANC contacts. The World Health Organization (WHO) recommends 8 ANC contacts as of 2016; however, this recommendation was adopted at different times across countries, including Kenya. Kenya adopted the ANC8+ recommendation in 2022; all pregnancies before 2022 were still managed under the ANC4+ policy. Since the survey included births in the five years preceding the survey, we used ANC4+ as the most appropriate indicator for comparability during the study period.

ANC uptake was coded as binary (0 for fewer than 4 ANC visits; 1 for 4 or more ANC visits). Skilled birth attendance was defined as delivery assisted by qualified healthcare personnel (midwife, nurse, auxiliary midwife, or doctor) and coded as binary (0 = no; 1 = yes). Postnatal care attendance was defined as receipt of care within 48 hours of delivery and coded as binary (0 = no; 1 = yes). The indicators were combined into a multinomial outcome variable with three categories: no continuum (none of the three services received), partial continuum (at least one but not all services received), and complete continuum (all three services received). Previous studies have used similar analytical approaches to assess maternal healthcare utilization [17, 18].

### Explanatory Variables

We included sociodemographic, household, reproductive, and media exposure factors as independent variables. Sociodemographic factors included age in 5-year groups, marital status, occupation, and education level. Household characteristics included household size, wealth index, sex of the household head, and place of residence. Reproductive factors included birth order and pregnancy intention. Media exposure factors included the frequency of reading newspapers, watching television, and listening to the radio.

### Data Management and Statistical Analysis

We used Stata version 16.0 (Stata Corp, College Station, TX) for analysis. Data cleaning and management were performed to prepare the dataset for analysis. Sampling weights provided by the DHS were applied to correct for over- and under-sampling. The survey’s complexity was addressed by clustering at the primary sampling unit (PSU) level and by stratification. Descriptive statistics summarized participant characteristics. We presented categorical variables as weighted frequencies and percentages. We estimated the prevalence of the continuum of maternal healthcare utilization using survey-weighted proportions. Bivariate analysis was conducted using the design-based Pearson chi-square. Variables with a p-value <0.20 in the bivariate analysis, along with those identified in prior literature as determinants of maternal healthcare utilization, were considered for inclusion in the multivariable model. We assessed multicollinearity among independent variables using the Variance Inflation Factor (VIF), with values>10 indicating multicollinearity. No evidence of multicollinearity was observed (VIF range: 1.06-5.10).

A survey-weighted multinomial logistic regression was fitted to assess inequalities in the continuum of maternal healthcare utilization. The outcome variable had three categories: no continuum, partial continuum, and complete continuum, with no continuum used as the reference category. Significant variables from the bivariate analysis were entered simultaneously into the model. Results were presented as adjusted relative risk ratios (aRRR) with 95% confidence intervals (CIs). Statistical significance was assessed at p < 0.05.

## Results

### Descriptive Characteristics

A weighted sample of 13,612 women who had a live birth within 5 years preceding the survey was included in the analysis. Overall, 67% of women had four or more ANC contacts, 62% had Skilled birth attendance, and 54% received postnatal care within 48 hours of delivery. (See **Table 1**). Most women were aged 25-29 years (29%), and a majority were in union (77.6%). More than half (55.1%) had attained secondary or higher education. Approximately 62% of the respondents were employed, and 61.6% were from households with five or fewer members. In terms of socioeconomic status, 23.4% of women were in the richest quintile, while 19.5% were in the poorest. Nearly half (46.6%) had a birth order greater than two, and 45.0% reported their most recent pregnancy as planned. Regarding media exposure, 53.5% watched television at least once per week, 62.6% reported listening to the radio at least once per week, and 82.7% reported not reading newspapers at all. The majority (60.7%) resided in the rural areas.

**Table 1:** Characteristics of Women in the Study.

| Variable Category | N=13612 |
| --- | --- |
| Age in Years |  |
| 15-19 | 732(5.4) |
| 20-24 | 3160(23.2) |
| 25-29 | 3948(29.0) |
| 30-34 | 2773(20.4) |
| 35-39 | 1993(14.6) |
| 40-44 | 797(5.9) |
| 45-49 | 209(1.5) |
| **Marital Status** |  |
| Not in Union | 3042(22.4) |
| In Union | 10570(77.6) |
| **Level of Education** |  |
| No education | 1097(8.1) |
| Primary Education | 5017(36.8) |
| Secondary or Higher | 7498(55.1) |
| **Occupation** |  |
| Unemployed | 5141(37.8) |
| Employed | 8471(62.2) |
| **Family Size** |  |
| <= 5 Members | 8380(61.6) |
| > 5 Members | 5232(38.4) |
| **HH Head is a Woman** |  |
| No | 9499(69.8) |
| Yes | 4113(30.2) |
| **Wealth Index** |  |
| Poorest | 2659(19.5) |
| Poorer | 2401(17.6) |
| Middle | 2421(17.8) |
| Richer | 2953(21.7) |
| Richest | 3178(23.4) |
| **Birth Order** |  |
| 1 | 3973(29.1) |
| 2 | 3301(24.3) |
| >2 | 6338(46.6) |
| **Planned pregnancy** |  |
| No | 7493(55.0) |
| Yes | 6119(45.0) |
| **Reads Newspaper** |  |
| Not at all | 11260(82.7) |
| Less than once per week | 1415(10.4) |
| At least once per week | 937(6.9) |
| **Watch TV** |  |
| Not at all | 4964(36.5) |
| Less than once per week | 1359(10.0) |
| At least once per week | 7289(53.5) |
| **Listen to Radio** |  |
| Not at all | 3314(24.4) |
| Less than once per week | 1775(13.0) |
| At least once per week | 8523(62.6) |
| **Residence** |  |
| Urban | 5345(39.3) |
| Rural | 8267(60.7) |

### Prevalence of Continuum of Maternal Healthcare Utilization

Overall, 60.8% (95% CI: 59.3 - 62.2%) of women experienced a partial continuum of care, 36.6% (95% CI: 35.1 - 38.0) achieved a complete continuum of care, and 2.7% (95% CI: 2.37 - 3.02) had no continuum of care. See **Fig 1**.

**Fig 1.**
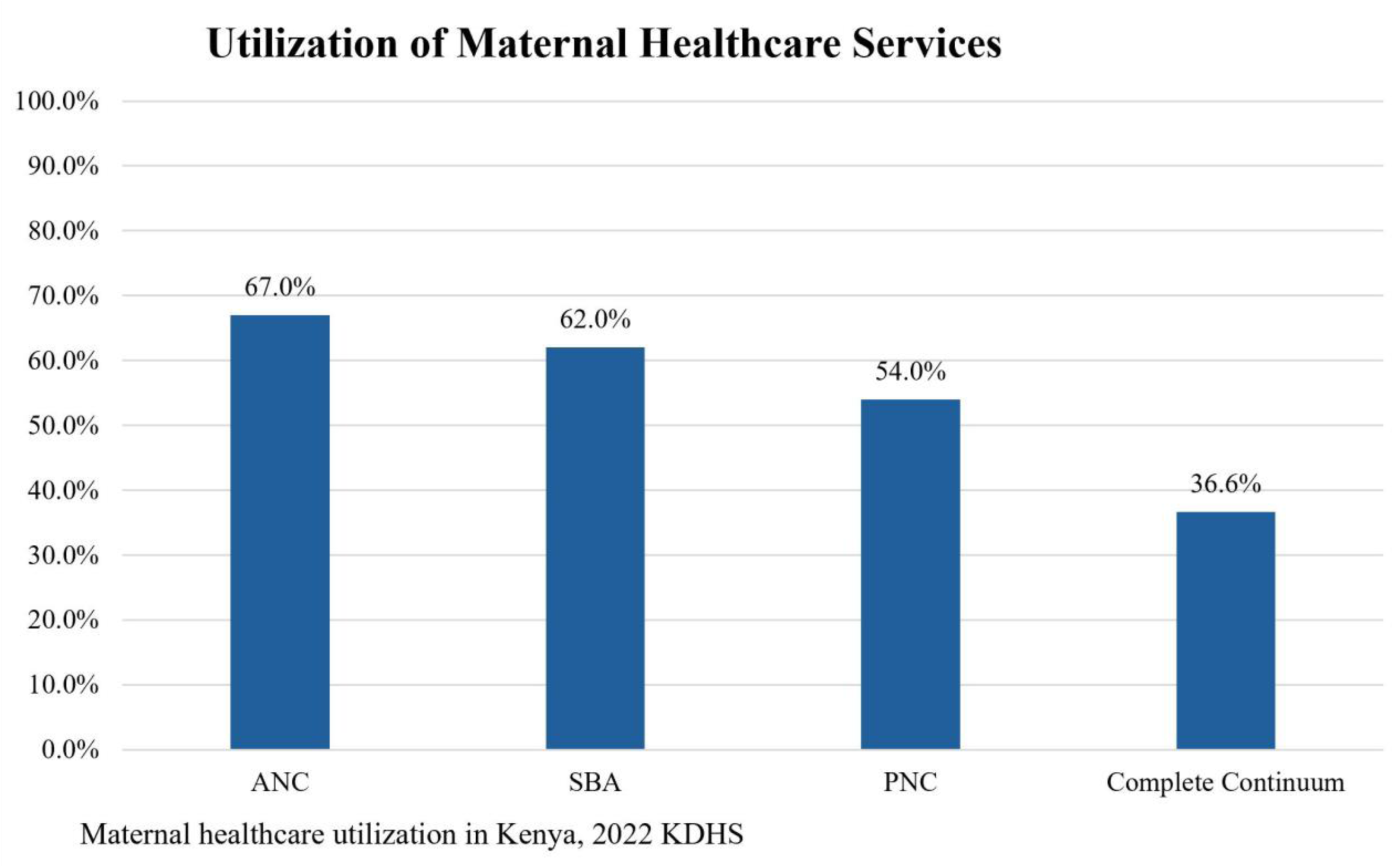
Maternal Healthcare Utilization and Continuum of Care.

### Bivariate analysis

We did a bivariate analysis, and all the variables were significantly associated with the continuum of maternal healthcare utilization. These variables were then inserted into the multivariable multinomial model. (See **Table 2**)

**Table 2:**
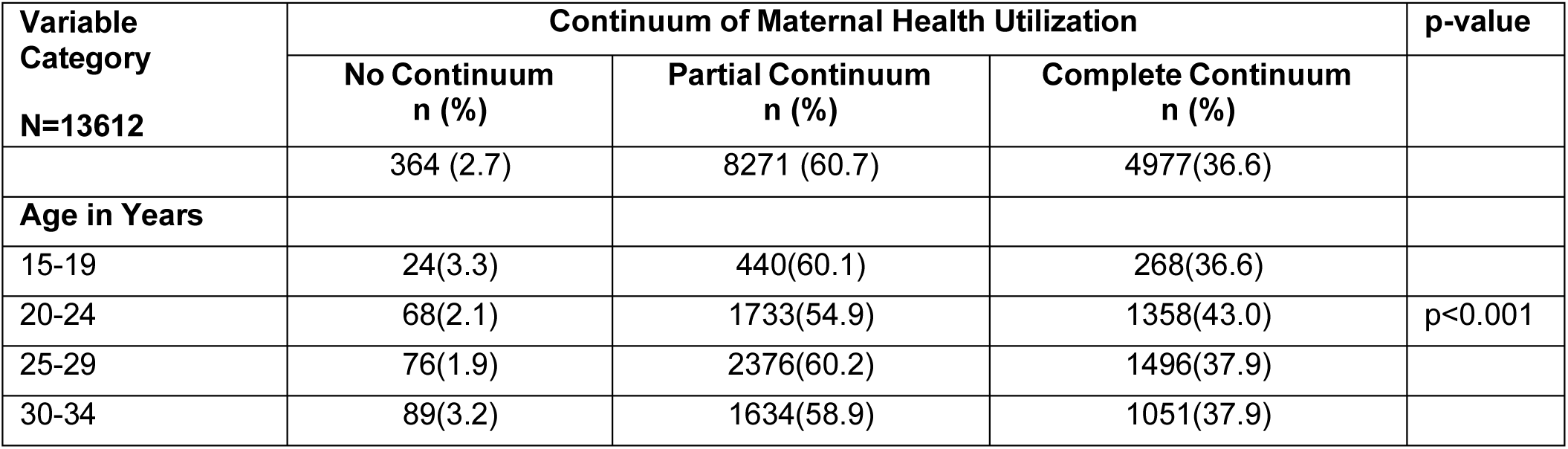
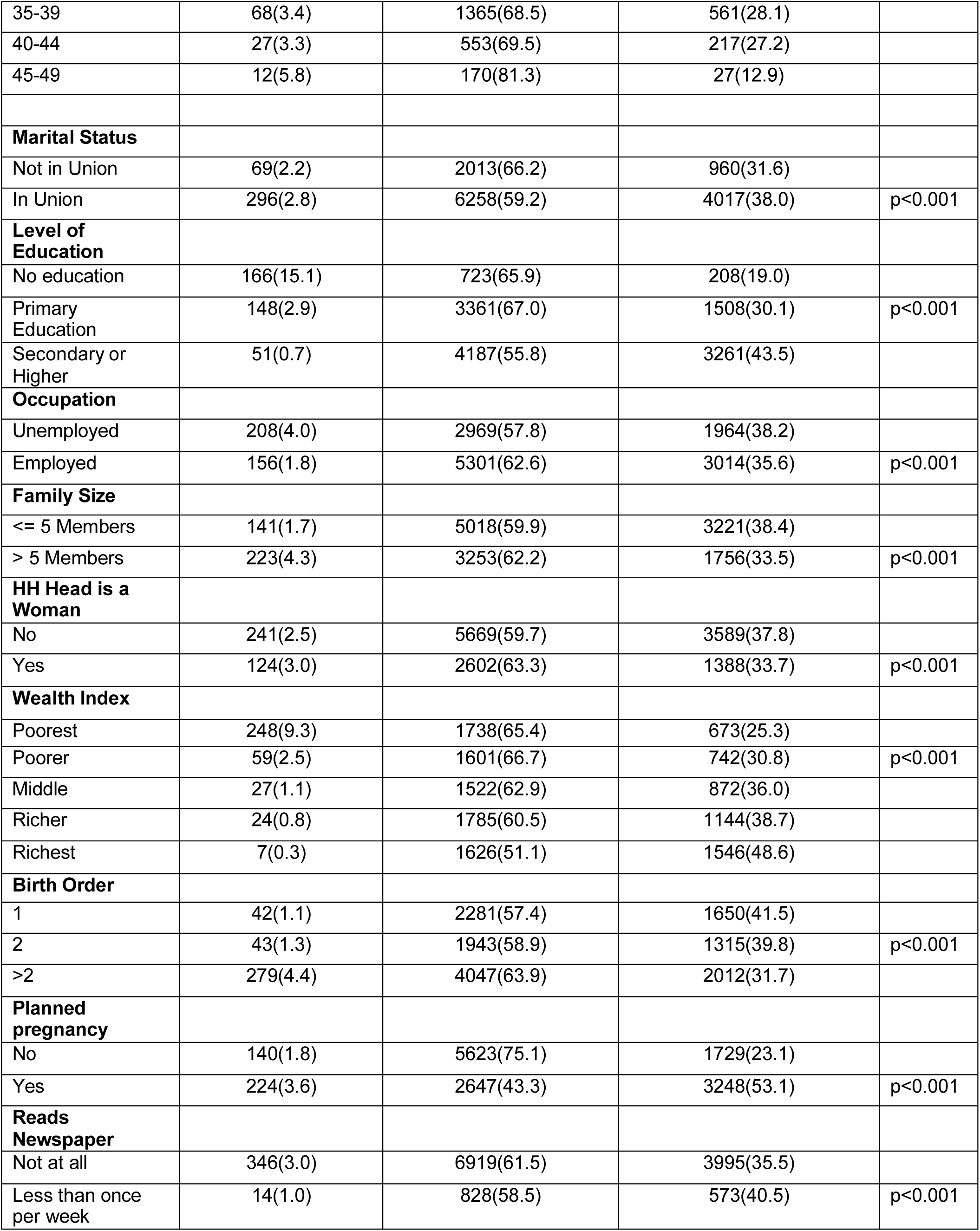
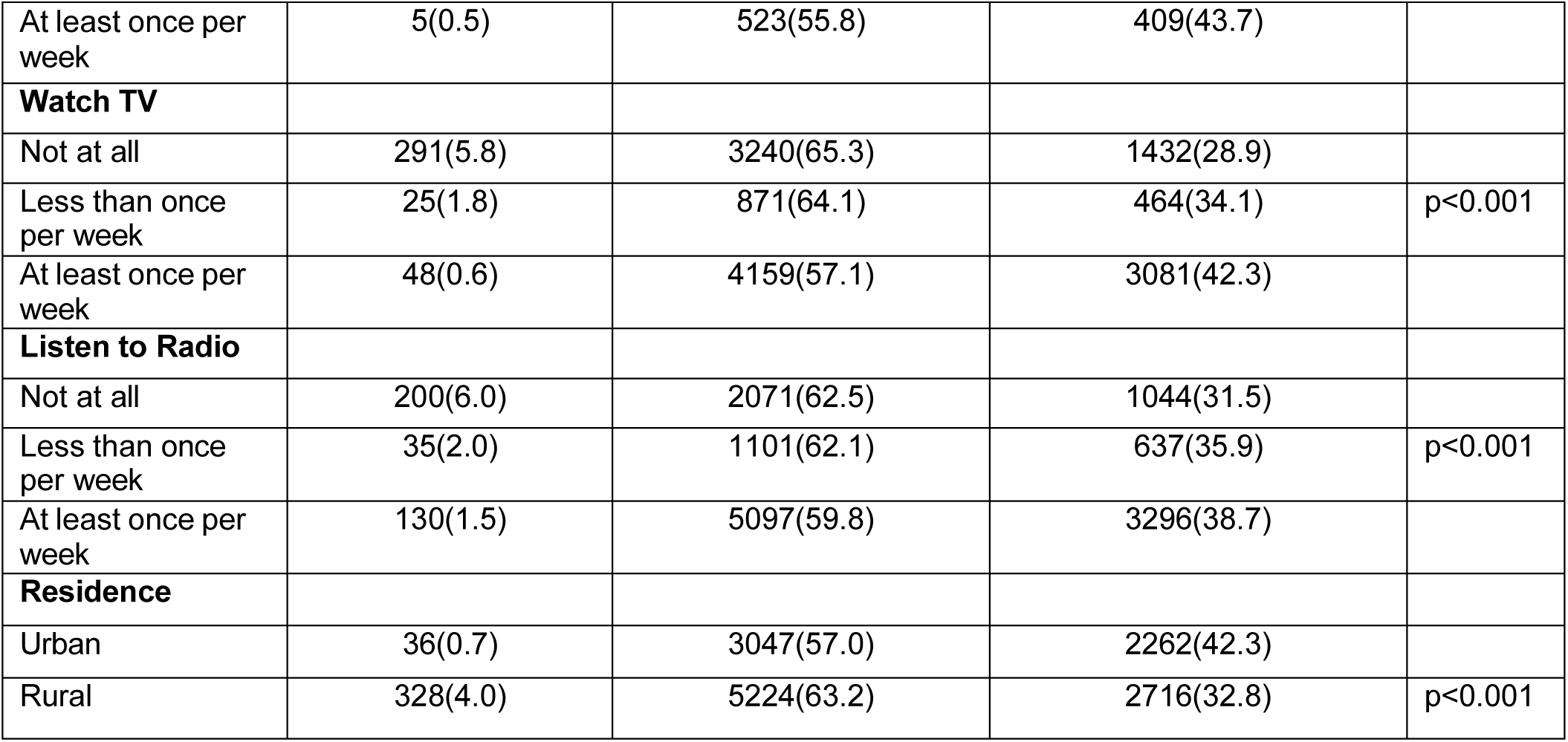
Bivariate Multinomial Regression.

### Multinomial regression analysis

A survey-weighted multinomial logistic regression was fitted to identify the inequalities in the continuum of maternal healthcare utilization, with no continuum as the reference category.

Women aged 25-29 years had a significantly higher likelihood of both partial (aRRR = 3.59; 95% CI: 2.08–6.19) and complete continuum (aRRR = 1.87; 95% CI: 1.08–3.25) than those aged 15–19 years, as seen in **Table 3**. Women in union were more likely to attain a complete continuum (aRRR = 1.50; 95% CI: 1.05–2.14). Education showed a strong positive gradient. Women with secondary education or higher had a significantly higher likelihood of both complete continuum (aRRR = 8.49; 95% CI: 4.91–14.66) and partial continuum (aRRR = 3.03; 95% CI: 1.79–5.14) than those with no formal education. Similarly, women with primary education had an increased likelihood of partial (aRRR = 2.06; 95% CI: 1.53-2.77) and complete continuum (aRRR = 4.12; 95% CI: 2.91-5.82) than those with no formal education.

**Table 3:** Determinants of Continuum of Maternal Healthcare Utilization in Kenya.

| Variable | Partial continuum<br>aRRR (95% CI) | p-value | Complete Continuum<br>aRRR (95% CI) | p-value |
| --- | --- | --- | --- | --- |
| **Age in Years** |  |  |  |  |
| 15-19 | Ref |  | Ref |  |
| 20-24 | 1.69 (1.01–2.85) * | 0.048 | 1.40 (0.82–2.38) | 0.213 |
| 25-29 | 3.59 (2.08–6.19) ** | 0.000 | 1.87 (1.08–3.25) * | 0.026 |
| 30-34 | 3.24 (1.82–5.74) ** | 0.000 | 1.75 (0.98–3.14) | 0.058 |
| 35-39 | 3.90 (2.12–7.16) ** | 0.000 | 1.42 (0.77–2.64) | 0.262 |
| 40-44 | 3.94 (2.07–7.49) ** | 0.000 | 1.55 (0.79–3.05) | 0.202 |
| 45-49 | 3.64 (1.62–8.17) * | 0.002 | 0.73 (0.28–1.95) | 0.533 |
| **Marital Status** |  |  |  |  |
| Not in Union | Ref |  | Ref |  |
| In a Union | 1.36 (0.96–1.91) | 0.081 | 1.5 (1.05–2.14) * | 0.027 |
| **Education level** |  |  |  |  |
| No Education | Ref |  | Ref |  |
| Primary Education | 2.06 (1.53–2.77) ** | 0.000 | 4.12 (2.91–5.82) ** | 0.000 |
| Secondary Education or Higher | 3.03 (1.79–5.14) ** | 0.000 | 8.49 (4.91–14.66) ** | 0.000 |
| **Occupation** |  |  |  |  |
| Not Employed | Ref |  | Ref |  |
| Employed | 1.09 (0.86–1.38) | 0.487 | 0.92 (0.72–1.19) | 0.530 |
| **Family Size** |  |  |  |  |
| **<=5 Members** | Ref |  | Ref |  |
| > 5 Members | 0.79 (0.60–1.03) | 0.080 | 0.95 (0.72–1.26) | 0.736 |
| **Woman is the Household head** |  |  |  |  |
| No | Ref |  | Ref |  |
| Yes | 1.05 (0.82–1.35) | 0.699 | 1.14 (0.87–1.48) | 0.344 |
| **Wealth Index** |  |  |  |  |
| Poorest | Ref |  | Ref |  |
| Poorer | 1.74 (1.24–2.44) * | 0.001 | 2 (1.40–2.85) ** | 0.000 |
| Middle | 2.66 (1.57–4.48) ** | 0.000 | 3.49 (2.06–5.92) ** | 0.000 |
| Richer | 2.05 (1.05–4.01) * | 0.036 | 3.05 (1.53–6.07) * | 0.002 |
| Richest | 4.14 (1.25–13.71) * | 0.020 | 9.38 (2.80–31.44) ** | 0.000 |
| **Birth Order** |  |  |  |  |
| 1 | Ref |  | Ref |  |
| 2 | 0.64 (0.41–0.99) * | 0.046 | 0.64 (0.41–1.00) | 0.053 |
| >2 | 0.26 (0.17–0.42) ** | 0.000 | 0.32 (0.20–0.51) ** | 0.000 |
| **Planned pregnancy** |  |  |  |  |
| No | Ref |  | Ref |  |
| Yes | 0.42 (0.33–0.53) ** | 0.000 | 1.79 (1.39–2.32) ** | 0.000 |
| **Read Newspaper** |  |  |  |  |
| Not at all | Ref |  |  |  |
| Less than once a week | 1.19 (0.60–2.35) | 0.619 | 1.28 (0.65–2.54) | 0.479 |
| At least once a week | 1.46 (0.60–3.58) | 0.407 | 1.46 (0.60–3.60) | 0.406 |
| **Watch TV** |  |  |  |  |
| Not at all | Ref |  | Ref |  |
| Less than once a week | 1.2 (0.76–1.88) | 0.437 | 1.30 (0.82–2.07) | 0.271 |
| At least once a week | 1.7 (1.09–2.65) * | 0.018 | 1.84 (1.18–2.87) * | 0.008 |
| **Listen radio** |  |  |  |  |
| Not at all | Ref |  | Ref |  |
| Less than once a week | 1.54 (0.99–2.39) | 0.055 | 1.68 (1.07–2.66) * | 0.025 |
| At least once a week | 1.38 (1.05–1.81) * | 0.020 | 1.59 (1.19–2.12) * | 0.002 |
| **Residence** |  |  |  |  |
| Urban | Ref |  | Ref |  |
| Rural | 0.57 (0.33–0.96) * | 0.035 | 0.61 (0.36–1.06) | 0.081 |

\*p<0.05; \*\* p<0.001, aRRR – adjusted Relative Risk Ratio

A strong wealth gradient was observed. Compared with women in the poorest households, those in the richest quintile had a higher likelihood of complete continuum (aRRR = 9.38; 95% CI: 2.80–31.44) and partial continuum (aRRR = 4.14; 95% CI: 1.25-13.71). Higher birth order was negatively associated with the continuum of care. Women with a birth order greater than two had a reduced likelihood of complete continuum (aRRR = 0.32; 95% CI: 0.20–0.51) and partial continuum (aRRR = 0.26; 95% CI: 0.17-0.42). Women with planned pregnancies had an increased likelihood of complete continuum (aRRR = 1.79; 95% CI: 1.39–2.32) and a lower likelihood of partial continuum (aRRR = 0.42; 95% CI: 0.33-0.53).

Media exposure was significantly associated with improved continuum. Women who watched television at least once a week had a higher likelihood of both partial (aRRR = 1.7; 95% CI: 1.09-2.65) and complete (aRRR = 1.84; 95% CI: 1.18-2.87) continuum. Similarly, listening to the radio at least once a week increased the likelihood of partial (aRRR = 1.38; 95% CI: 1.05-1.81) and complete (aRRR = 1.59; 95% CI: 1.19-2.12) continuum. Women residing in rural areas had a lower likelihood of partial continuum (aRRR = 0.57; 95% CI: 0.33-0.96).

## Discussion

This study explored the inequalities in the continuum of maternal healthcare utilization in Kenya using a nationally representative survey. To our knowledge, this study is the first to assess the full spectrum of maternal healthcare utilization encompassing SBA, ANC, and PNC in Kenya. Prior studies have assessed these components individually and not as a composite measure. The analysis revealed high individual service utilization and very low completion of the continuum (36.6%). Most of the women (60.8%) attained a partial continuum of care, indicating substantial drop-offs across the various stages of care. This pattern is seen in prior studies in SSA literature that demonstrate discontinuities in access to ANC, skilled delivery, and postnatal care [4, 13, 19]. These drop-offs indicate systemic gaps in service integration and continuity.

### Education & Wealth Inequalities

The likelihood of completing the continuum was higher among women from the richest quintile than among those from the poorest, and among those with higher education than among those with no education. This is consistent with prior studies conducted in SSA [12], India [18] and Afghanistan [20] that demonstrate that wealth and education are among the most consistent predictors of maternal healthcare utilization. Education enhances health literacy, decision-making capacity, and autonomy [21, 22], while wealth minimizes the logistical and financial barriers to accessing care [11, 12, 23]. Despite Kenya’s introduction of free maternity services (*Linda Mama program*) across all public facilities in 2013 to reduce financial barriers to accessing skilled delivery and maternal care, wealth disparities persist. The persistence in the wealth gradient in our analysis suggests that financial barriers alone do not explain the inequalities in the completion of the continuum. There are indirect costs, such as transportation, and out-of-pocket charges for ancillary services, that disadvantage women from poorer households. Additionally, the availability and quality of services in facilities that predominantly serve women from poorer households may also contribute to the dropout in the various stages of care.

### Age and Marital Union differences

Women’s age affected continuity of care, with those aged 25-29 years more likely to achieve both partial and complete continuum than adolescents. This suggests greater engagement with healthcare services during peak reproductive years. Older women, on the other hand, were more likely to attain a partial continuum but not a complete continuum. This is likely due to challenges of competing responsibilities, a reduced perceived need for care, and prior childbirth experience [24]. Women in union also had a higher likelihood of completion of the continuum, emphasizing that partner involvement and support play an important role in facilitating access to maternal healthcare services. This is evidenced by previous studies that highlight the importance of male partner involvement in the use of maternal healthcare services [25].

### Reproductive factors

Parity was an important predictor of discontinuity. Women with a higher birth order were less likely to achieve a partial or complete continuum. This aligns with existing literature showing an inverse relationship between maternal healthcare utilization and parity. In a study by Larsen et al. [24], results showed that women with a higher birth order were 84% less likely to utilize maternal healthcare services as compared to women with a lower birth order. This could be explained by the fact that multiparous women often face competing household responsibilities and rely on previous birth experience, leading to reduced engagement with healthcare providers during their pregnancies [26–28]. Additionally, some who have experienced high service costs and poor attitudes from healthcare professionals during previous pregnancies become dissatisfied with healthcare services, leading to reduced utilization, as seen in previous studies [29–31]. Planned pregnancies also revealed a distinct pattern in the continuum of care. This is consistent with Dibaba et al., who found that pregnancy intention was associated with increased maternal healthcare utilization [32]. Women with planned pregnancies had a higher likelihood of complete continuum and a lower likelihood of partial continuum. This is because planned pregnancies are associated with more consistent and intentional health-seeking behaviors, leading to the completion of the full continuum of care rather than discontinuity in care.

### Geographic disparities

Geographic inequalities were also observed. Women residing in rural areas had a lower likelihood of achieving partial continuum, consistent with other studies that reported lower utilization of maternal healthcare services among rural women compared with their urban counterparts [10, 23, 33–35]. There is a spatial concentration of geographic inequalities in Kenya, especially in the northern, eastern, and coastal areas, as well as parts of western Kenya. Low rates of maternal utilization have been observed in these areas in previous rounds of KDHS. This could be explained by long distances and rugged terrain, which make access to facilities cumbersome for pregnant and postpartum women [12, 34]. Additionally, in many rural areas, there are fewer facilities, most of which are not fully equipped and have limited healthcare providers, which limit access and the quality of maternal healthcare services, leading to reduced utilization [34].

### Media Exposure

Media exposure was positively associated with a continuum of maternal healthcare utilization, a finding supported by several studies conducted in other low- to middle-income countries [36, 37]. Television and radio specifically were associated with a higher likelihood of both partial and complete continuum. These channels play an important role in relaying health information, such as the importance of antenatal care services, exposure to postpartum care, safe delivery practices, and even danger signs during pregnancy [36]. They help women understand the importance of seeking timely care and influence their health-seeking behaviors, thereby increasing utilization of these services. [37].

## Strengths and Limitations

This study has several notable strengths and limitations. To begin with, this study utilizes data from the nationally representative 2022 KDHS, which was collected through standardized measures and tools, making it generalizable to women in Kenya. Additionally, previous studies in Kenya have assessed maternal utilization using these services independently. However, this study used a composite measure of three key maternal health services to assess the quality of care. Moreover, a multinomial regression model assessing no, partial, and complete categories enabled simultaneous comparison across categories, thereby providing more insights than a binary model.

Despite the strengths, the study had a few limitations. The data is based on self-reported information over a 5-year period. This may be subject to recall bias, especially for services such as antenatal visits and postnatal care. Additionally, this study is a cross-sectional survey; thus, establishing causal relationships between the explanatory variables and the continuum of maternal healthcare utilization may be difficult. Despite these limitations, the study provides population-level evidence of inequalities in maternal healthcare utilization and offers valuable insights into programmatic and policy interventions in Kenya.

## Conclusion

Continuum of maternal healthcare utilization remains suboptimal in Kenya, with substantial gaps in the continuity of care despite a relatively high engagement with individual services. This study identified important socioeconomic and sociodemographic inequalities, including maternal age, parity, education, wealth quintile, pregnancy intention, residence, and media exposure, that were associated with both partial and complete care in the continuum of care. The government and other stakeholders should integrate financial policies with community-based interventions, such as targeted health education programs and community health worker outreach, to ensure continuity of care. Healthcare programs and providers should expand access to family planning services and promote male partner involvement to increase utilization. Finally, stakeholders, the private sector, and health promotion departments in the Ministry of Health should increase media sensitization to promote maternal health education initiatives.

## Data Availability

All data produced is available online. Data access was granted by the DHS program (https://www.dhsprogram.com/) upon a formal request.

## Ethical Statement and Data Availability

The original survey obtained ethical approval from the Kenya Medical Research Institute (KEMRI) Scientific and Ethics Review Unit (SERU) and the ICF Institutional Review Board (IRB). All participants provided informed consent before data collection. For this study, we obtained ethical approval from the American University of Antigua (AUA) IRB (Approval No: AUAIRB26004). The data set obtained contains no personally identifiable information, as it was de-identified before being made publicly accessible. Data access was granted by the DHS program (https://www.dhsprogram.com/) upon a formal request.

## Acknowledgement

We want to acknowledge the Demographic Health Survey program for providing access to the data used for this study.

## References

1. WHO. Trends in maternal mortality 2000 to 2020: estimates by WHO, UNICEF, UNFPA, World Bank Group and UNDESA/Population Division. World Health Organization, 2023 9240068759.

2. Kerber KJ, de Graft-Johnson JE, Bhutta ZA, Okong P, Starrs A, Lawn JE. Continuum of care for maternal, newborn, and child health: from slogan to service delivery. Lancet. 2007;370(9595):1358–69.

3. Oyedele OK, Fagbamigbe AF, Akinyemi OJ, Adebowale AS. Coverage-level and predictors of maternity continuum of care in Nigeria: implications for maternal, newborn and child health programming. BMC Pregnancy Childbirth. 2023;23(1):36.

4. Singh K, Story WT, Moran AC. Assessing the continuum of care pathway for maternal health in South Asia and sub-Saharan Africa. Maternal and Child Health J. 2016;20(2):281–9.

5. Tessema ZT, Teshale AB, Tesema GA, Tamirat KS. Determinants of completing recommended antenatal care utilization in sub-Saharan from 2006 to 2018: evidence from 36 countries using Demographic and Health Surveys. BMC Pregnancy Childbirth. 2021;21(1):192.

6. Benova L, Owolabi O, Radovich E, Wong KL, Macleod D, Langlois EV, et al. Provision of postpartum care to women giving birth in health facilities in sub-Saharan Africa: A cross-sectional study using Demographic and Health Survey data from 33 countries. PLoS Med. 2019;16(10):e1002943.

7. Bhutta ZA, Das JK, Bahl R, Lawn JE, Salam RA, Paul VK, et al. Can available interventions end preventable deaths in mothers, newborn babies, and stillbirths, and at what cost? Lancet. 2014;384(9940):347–70.

8. Usman M, Banerjee A, Srivastava S. Association between maternal health continuum of care and child survival: evidence from a population based survey. Children and Youth Services Review. 2021;128:106134.

9. Zelka MA, Yalew AW, Debelew GT. The effects of completion of continuum of care in maternal health services on adverse birth outcomes in Northwestern Ethiopia: a prospective follow-up study. Reprod Health. 2022;19(1):200.

10. Bobo FT, Asante A, Woldie M, Dawson A, Hayen A. Spatial patterns and inequalities in skilled birth attendance and caesarean delivery in sub-Saharan Africa. BMJ Global Health. 2021;6(10).

11. Boerma T, Requejo J, Victora CG, Amouzou A, George A, Agyepong I, et al. Countdown to 2030: tracking progress towards universal coverage for reproductive, maternal, newborn, and child health. Lancet. 2018;391(10129):1538–48.

12. Samuel O, Zewotir T, North D. Decomposing the urban–rural inequalities in the utilisation of maternal health care services: evidence from 27 selected countries in Sub-Saharan Africa. Reprod Health. 2021;18(1):216.

13. Goli S, Nawal D, Rammohan A, Sekher T, Singh D. Decomposing the socioeconomic inequality in utilization of maternal health care services in selected countries of South Asia and sub-Saharan Africa. J Biosoc Sci. 2018;50(6):749–69.

14. Kruk ME, Gage AD, Arsenault C, Jordan K, Leslie HH, Roder-DeWan S, et al. High-quality health systems in the Sustainable Development Goals era: time for a revolution. The Lancet Global Health. 2018;6(11):e1196–e252.

15. KNBS and ICF. Kenya Demographic and Health Survey 2022. Nairobi, Kenya, and Rockville, Maryland, USA: KNBS and ICF, 2023.

16. Afulani PA, Buback L, Essandoh F, Kinyua J, Kirumbi L, Cohen CR. Quality of antenatal care and associated factors in a rural county in Kenya: an assessment of service provision and experience dimensions. BMC Health Serv Res. 2019;19(1):684.

17. Adedokun ST, Uthman OA, Bisiriyu LA. Determinants of partial and adequate maternal health services utilization in Nigeria: analysis of cross-sectional survey. BMC Pregnancy Childbirth. 2023;23(1):457.

18. Gandhi S, Gandhi S, Dash U, Suresh Babu M. Predictors of the utilisation of continuum of maternal health care services in India. BMC Health Serv Res. 2022;22(1):602.

19. Mohan D, LeFevre AE, George A, Mpembeni R, Bazant E, Rusibamayila N, et al. Analysis of dropout across the continuum of maternal health care in Tanzania: findings from a cross-sectional household survey. Health Policy Plan. 2017;32(6):791–9.

20. Mumtaz S, Bahk J, Khang Y-H. Current status and determinants of maternal healthcare utilization in Afghanistan: Analysis from Afghanistan Demographic and Health Survey 2015. PLoS One. 2019;14(6):e0217827.

21. Gebeyehu NA, Gelaw KA, Lake EA, Adela GA, Tegegne KD, Shewangashaw NE. Women decision-making autonomy on maternal health service and associated factors in low-and middle-income countries: Systematic review and meta-analysis. Womens Health. 2022;18:17455057221122618.

22. Idris IB, Hamis AA, Bukhori ABM, Hoong DCC, Yusop H, Shaharuddin MA-A, et al. Women’s autonomy in healthcare decision making: a systematic review. BMC Womens Health. 2023;23(1):643.

23. Tey N, Lai S. Correlates of and barriers to the utilization of health services for delivery in South Asia and Sub-Saharan Africa. Sci World J. 2013; 2013: 423403.

24. Larsen A, Exavery A, Phillips JF, Tani K, Kanté AM. Predictors of health care seeking behavior during pregnancy, delivery, and the postnatal period in rural Tanzania. Maternal and Child Health J. 2016;20(8):1726–34.

25. Yargawa J, Leonardi-Bee J. Male involvement and maternal health outcomes: systematic review and meta-analysis. Journal of Epidemiology and Community Health. 2015;69(6):604–12.

26. Bain LE, Aboagye RG, Dowou RK, Kongnyuy EJ, Memiah P, Amu H. Prevalence and determinants of maternal healthcare utilisation among young women in sub-Saharan Africa: cross-sectional analyses of demographic and health survey data. BMC Public Health. 2022;22(1):647.

27. Dimbuene ZT, Amo-Adjei J, Amugsi D, Mumah J, Izugbara CO, Beguy D. Women’s education and utilization of maternal health services in Africa: a multi-country and socioeconomic status analysis. J Biosoc Sci. 2018;50(6):725–48.

28. Tsawe M, Susuman AS. Determinants of access to and use of maternal health care services in the Eastern Cape, South Africa: a quantitative and qualitative investigation. BMC Res Notes. 2014;7(1):723.

29. Amu H, Nyarko SH. Satisfaction with maternal healthcare services in the Ketu South municipality, Ghana: a qualitative case study. BioMed Res Intl. 2019;2019(1):2516469.

30. Mannava P, Durrant K, Fisher J, Chersich M, Luchters S. Attitudes and behaviours of maternal health care providers in interactions with clients: a systematic review. Glob Health. 2015;11(1):36.

31. Matsuoka S, Aiga H, Rasmey LC, Rathavy T, Okitsu A. Perceived barriers to utilization of maternal health services in rural Cambodia. Health Policy. 2010;95(2-3):255–63.

32. Dibaba Y, Fantahun M, Hindin MJ. The effects of pregnancy intention on the use of antenatal care services: systematic review and meta-analysis. Reprod Health. 2013;10(1):50.

33. Gudu W, Addo B. Factors associated with utilization of skilled service delivery among women in rural Northern Ghana: a cross sectional study. BMC Pregnancy Childbirth. 2017;17(1):159.

34. Tanou M, Kamiya Y. Assessing the impact of geographical access to health facilities on maternal healthcare utilization: evidence from the Burkina Faso demographic and health survey 2010. BMC Public Health. 2019;19(1):838.

35. Zelalem Ayele D, Belayihun B, Teji K, Admassu Ayana D. Factors affecting utilization of maternal healthcare services in Kombolcha District, eastern Hararghe zone, Oromia regional state, eastern Ethiopia. Intl Schol Res Not. 2014;2014(1):917058.

36. Acharya D, Khanal V, Singh JK, Adhikari M, Gautam S. Impact of mass media on the utilization of antenatal care services among women of rural community in Nepal. BMC Res Notes. 2015;8(1):345.

37. Fatema K, Lariscy JT. Mass media exposure and maternal healthcare utilization in South Asia. SSM-Pop Health. 2020;11:100614.

